# Validation of the LINEAR deflation algorithm of the Midmark IQvitals Zone Vital Signs Monitor: improving patient comfort from quicker BP acquisition

**DOI:** 10.1101/2023.07.28.23293336

**Authors:** Bruce S. Alpert

**Affiliations:** Department of Pediatrics, University of Tennessee Health Sciences Center, Memphis, Tennessee, USA

## Abstract

**Objectives:** To assess the accuracy of the Midmark IQvitals^®^ Zone™ wireless vital signs monitor LINEAR deflation algorithm to the requirements of the ANSI/AAMI/ISO 81060-2 Standard and the British Hypertension Society (BHS) Protocol.

**Methods:** The Standard and BHS testing each call for ≥ 85 subjects with requirements for gender, blood pressure (BP), and arm circumference. The testers performing auscultation were blinded. Statistical calculations as per requirements were performed.

**Results:** The mean ± SD for the 81060-2 Criterion 1 were 1.98 ± 6.90 mmHg for systolic and 0.54 ± 5.79 mmHg for diastolic BP. The Criterion 2 SD values were 5.60 mmHg for systolic and 5.26 mmHg for diastolic BP. All of these values passed the Standard requirements. The overall BHS rating was AA.

**Conclusions:** The Midmark LINEAR algorithm is validated based on the results. Use of the algorithm results in shorter cuff deflation times, thus improving patient comfort. The LINEAR algorithm has multiple features to improve BP measurement accuracy in critical patient populations.

---

The estimation of BP by use of automated sphygmomanometers, usually programmed with oscillometric technology, has become the accepted clinical method [1]. These devices are in use in professional healthcare sites, as well as out-of-office (home) sites. Devices must be validated according to the requirements of the ANSI/AAMI/ISO 81060-2, 2018 Standard [2]. In certain world markets, the British Hypertension Society (BHS) Protocol [3] is still in use.

As manufacturers seek to improve automated devices, both hardware and software (algorithm) modernization has occurred. The update goals are to improve the accuracy of the BP readings and lessen patient discomfort. A shorter time of cuff deflation will lead to improved patient comfort, especially in populations with high BP, high heart rate, neurologic challenges, and pediatric patients. A shorter duration of BP measurement also results in fewer artifacts from subject motion. The Midmark LINEAR deflation algorithm is available as a clinical preferences option on the tested device.

The LINEAR algorithm uses a novel adaptive technique based on the patient’s heart rate and the mean arterial pressure (MAP) to determine the optimal deflation rate during BP measurement. When a lower heart rate is detected, the deflation rate is set to a low value to ensure the system acquires the required number of pulses for the LINEAR algorithm to determine the BP. When the heart rate is higher, the deflation rate is accelerated. With more pulses occurring in a shorter time span, the accelerated deflation rate still allows the algorithm to measure the requisite number of pulses within each pressure range to achieve an accurate BP determination.

Optimization of the deflation rate helps to ensure the least time of arm compression needed and minimizes motion opportunities by minimizing the duration of the BP measurement process for each patient. Based on the total cycle time for BP acquisition among subjects in this study and the STEP deflation study [4], the aggregate result among all subjects is a deflation time that is approximately 40% faster when using the LINEAR algorithm compared to the Midmark STEP algorithm.

The Midmark LINEAR deflation algorithm also uses a novel technique based on an individual’s heart rate and pulse amplitude to extrapolate pulses. This provides a mechanism to retain accurate measurement of BP in situations of underinflation or aberrant pulses caused by artifacts. With this novel method of pulse extrapolation, the system requires fewer reinflation cycles and reduces overall acquisition time to improve overall efficiency and patient comfort.

## Material and Methods

The validation testing was performed by the staff at Clinimark, LLC in Louisville, CO, USA. This testing site has extensive experience with performance of ISO 81060-2 and BHS requirements.

Eighty-five subjects were included in the final analyses. The subjects were seated in the Midmark 626 Barrier-Free^®^ Examination Chair which helps ensure that proper patient positioning is achieved [1,4]. The chair has features and accessories that allow for feet flat on the floor, back supported, and arm supported with the cuff at the level of the heart. A total of six different cuffs were included for testing, covering limb circumference ranges from 18-50 cm.

The protocols followed the requirements which included data for gender, BP values, and arm circumferences. In addition to device readings, there were two blinded observers performing auscultation and a third observer who recorded the BP values and ensured that the readings for each inflation/deflation cycle for the two observers did not differ by > 4 mmHg. To be included in the analysis, the overall “drift” of the BP could not exceed 12 mmHg for systolic BP (SDP) and 8 mmHg for diastolic BP (DBP). The same-arm sequential protocol [2] was followed. Written informed consent was obtained from each subject. The study was approved by the Salus Institutional Review Board.

## Analyses

The BP reading from each of the blinded observers was recorded by Observer 3. The two values were averaged. The sequence of cycles followed that described in the ISO 81060-2 Standard. The order of cycles was manual, device, manual, device, manual, device, and finally manual. The averages of the two manual readings prior to and following each device reading were calculated. Thus, for each subject there were three sets of manual/device comparisons. This bracketing helps control for any BP change over time, by using manual readings just before and just after each device reading. From these data, means ± SDs were calculated for use in Criterion 1 and Criterion 2 requirements [2]. The calculations for the BHS Protocol were also performed.

## Results

Error was defined as device reading minus the manual readings. The results of the ISO 81060-2 Standard analyses are shown in Table 1.

**Table 1.** ISO 81060-2 Standard test results.

| LINEAR<br>algorithm | Criterion 1<br>mean (mmHg) ± SD | Criterion 2<br>SD (mmHg) | Pass/Fail |
| --- | --- | --- | --- |
| Systolic | 1.98 ± 6.90 | 5.60 | Pass |
| Diastolic | 0.54 ± 5.97 | 5.26 | Pass |

The Standard requires Bland–Altman plots demonstrating the scatter of data points for both SBP and DBP readings (Figure 1a and 1b).

**Figure 1.**
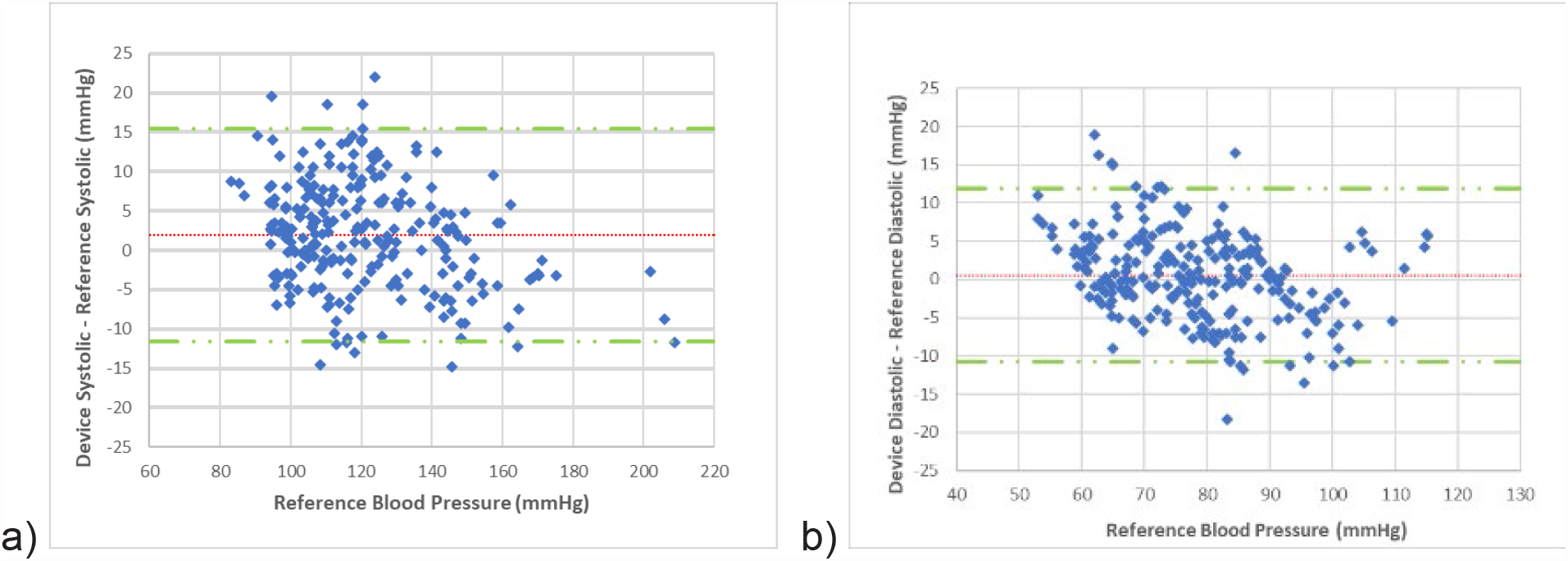
Bland–Altman plots demonstrating the scatter of data points for both (a) systolic and (b) diastolic blood pressure readings.

The data also were analyzed per the BHS Protocol. This includes “grades” from A-C for agreement between device and observer for the overall data set and for subscales using BP cut-off values. These results are shown in Table 2.

**Table 2.**
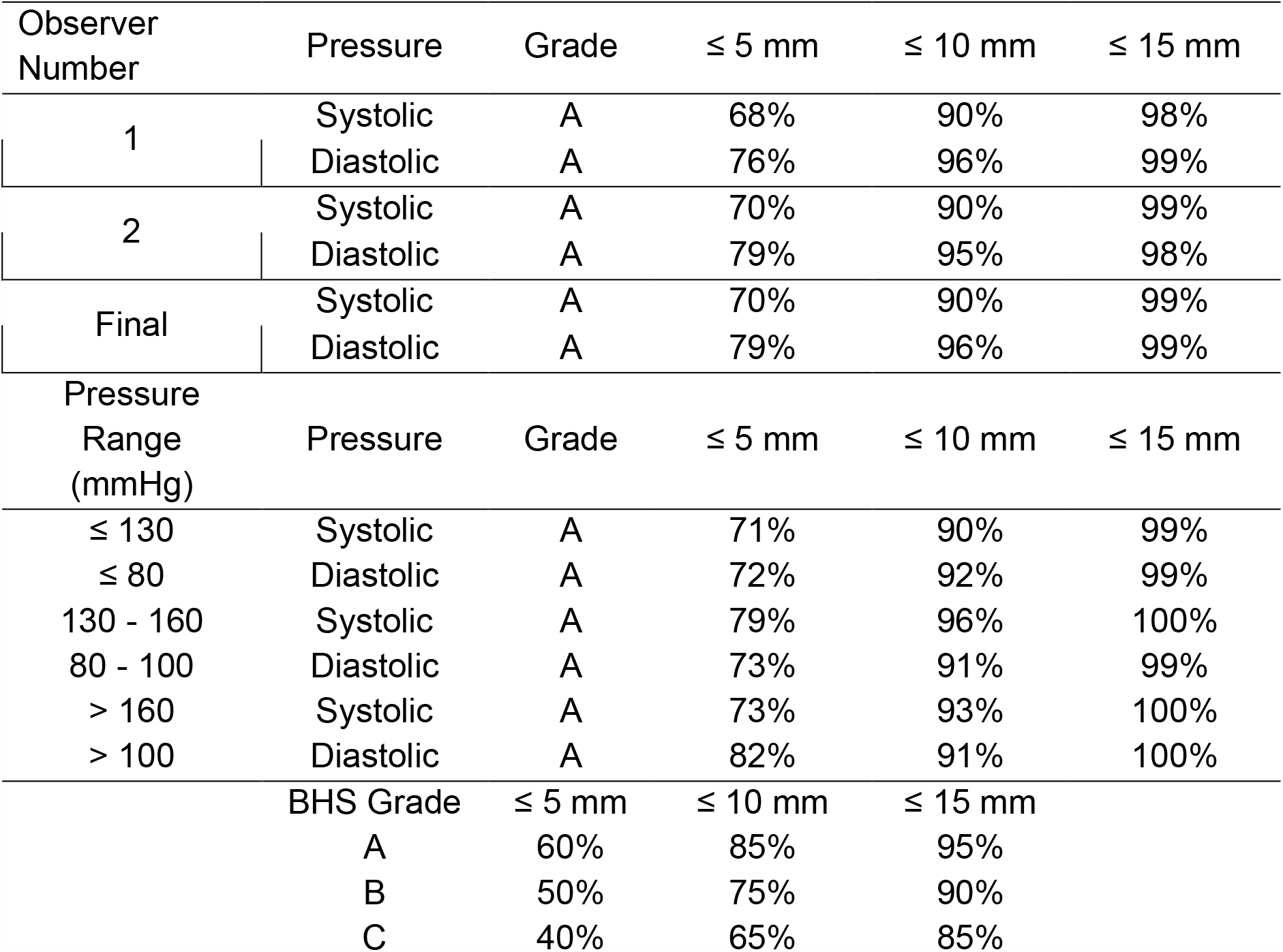
BHS Protocol assessment results.

| Observer Number | Pressure | Grade | ≤ 5 mm | ≤ 10 mm | ≤ 15 mm |
| --- | --- | --- | --- | --- | --- |
| 1 | Systolic | A | 68% | 90% | 98% |
|  | Diastolic | A | 76% | 96% | 99% |
| 2 | Systolic | A | 70% | 90% | 99% |
|  | Diastolic | A | 79% | 95% | 98% |
| Final | Systolic | A | 70% | 90% | 99% |
|  | Diastolic | A | 79% | 96% | 99% |
| Pressure Range (mmHg) | Pressure | Grade | ≤ 5 mm | ≤ 10 mm | ≤ 15 mm |
| ≤ 130 | Systolic | A | 71% | 90% | 99% |
| ≤ 80 | Diastolic | A | 72% | 92% | 99% |
| 130 - 160 | Systolic | A | 79% | 96% | 100% |
| 80 - 100 | Diastolic | A | 73% | 91% | 99% |
| > 160 | Systolic | A | 73% | 93% | 100% |
| > 100 | Diastolic | A | 82% | 91% | 100% |
|  | BHS Grade | ≤ 5 mm | ≤ 10 mm | ≤ 15 mm |  |
|  | A | 60% | 85% | 95% |  |
|  | B | 50% | 75% | 90% |  |
|  | C | 40% | 65% | 85% |  |

## Discussion

The IQvitals Zone with the LINEAR algorithm has novel features which will improve patient comfort. During inflation, the algorithm detects mean BP and limits inflation above SBP. The LINEAR deflation can be up to 10 mmHg per beat at slower heart rates. And, when compared to the algorithm used for STEP deflation, the LINEAR algorithm captures BP values in approximately 40% less time. Together, these improvements enhance patient comfort by limiting maximal inflation and speeding determination of DBP. This is especially beneficial in patients who have difficulty holding still during inflation/deflation cycles, such as individuals with neurologic disorders or children.

The LINEAR algorithm passed all requirements of the ISO 81060-2 Standard and obtained the highest grading per the BHS Protocol.

## Data Availability

All data produced in the present study are available upon reasonable request to the author.

